# Assessing the impact of data aggregation in model predictions of HAT transmission and control activities

**DOI:** 10.1101/19005991

**Authors:** M. Soledad Castaño, Martial L. Ndeffo-Mbah, Kat S. Rock, Cody Palmer, Edward Knock, Erick Mwamba Miaka, Joseph M. Ndung’u, Steve Torr, Paul Verlé, Simon E.F. Spencer, Alison Galvani, Caitlin Bever, Matt J. Keeling, Nakul Chitnis

**Author notes:** (MSC).

## Abstract

Since the turn of the century, the global community has made great progress towards the elimination of gambiense human African trypanosomiasis (HAT). Elimination programs, primarily relying on screening and treatment campaigns, have also created a rich database of HAT epidemiology. Mathematical models calibrated with these data can help to fill remaining gaps in our understanding of HAT transmission dynamics, including key operational research questions such as whether integrating vector control with current intervention strategies is needed to achieve HAT elimination. Here we explore, via an ensemble of models and simulation studies, which aspects of the available data and level of data aggregation, such as separation by disease stage, would be most useful for better understanding transmission dynamics and improving model reliability in making future predictions of control and elimination strategies.

**Author summary:** Human African tryposonomiasis (HAT), also known as sleeping sickness, is a parasitic disease with over 65 million people estimated to be living at risk of infection. Sleeping sickness consists of two stages: the first one is relatively mild but the second stage is usually fatal if untreated. The World Health Organization has targeted HAT for elimination as a public health problem by 2020 and for elimination of transmission by 2030. Regular monitoring updates indicate that 2020 elimination goals are likely to be achieved. This monitoring relies mainly on case report data that is collected through medical-based control activities — the main strategy employed so far in HAT control. This epidemiological data are also used to calibrate mathematical models that can be used to analyse current interventions and provide projections of potential intensified strategies.

We investigated the role of the type and level of aggregation of this HAT case data on model calibrations and projections. We highlight that the lack of detailed epidemiological information, such as missing stage of disease or truncated time series data, impacts model recommendations for strategy choice: it can misrepresent the underlying HAT epidemiology (for example, the ratio of stage 1 to stage 2 cases) and increase uncertainty in predictions. Consistently including new data from control activities as well as enriching it through cross-sectional (e.g. demographic or behavioural data) and geo-located data is likely to improve modelling accuracy to support planning, monitoring and adapting HAT interventions.

## Introduction

Human African trypanosomiasis (HAT) is a neglected tropical disease that affects people in resource-limited settings in sub-Saharan Africa, with more than 65 million people living at risk [1]. HAT is caused by a protozoan parasite and is transmitted between humans by biting tsetse flies. The gambiense form of the disease, caused by *Trypanosoma brucei gambiense*, is responsible for over 95% of human cases. This chronic disease progresses through two stages. The first stage can last for several years with relatively minor symptoms such as fever and headaches. Second stage patients show neuropsychiatric disorders (including sleep disturbances that led to the common name, sleeping sickness) and this stage is usually fatal without treatment. Currently available treatments are stage-dependent and so assessment of a patient’s stage - by analysing the cerebrospinal fluid for parasites and number of white blood cells - is a prerequisite for appropriate treatment.

Since the start of the 21^st^ Century, control activities against gambiense HAT have had a substantial impact on reducing disease transmission and burden in the main endemic regions [2]. These control efforts have raised expectations that elimination of gambiense HAT may be achievable [1, 3]. The World Health Organization (WHO) has therefore set indicators that target elimination of transmission (EOT) by 2030.

Although there were only 953 cases reported globally in 2018 [4], persistent foci of disease transmission remain a potential challenge for achieving the EOT goal. The Democratic Republic of Congo (DRC) has suffered from persistent infection, contributing between 78–91% of all global cases since 2010 [4].

Efforts to control HAT have mainly relied on screening, testing and treating the human population using active and/or passive surveillance. This has been the only intervention applied at large scale, and it seems likely that this is largely responsible for the precipitous decline in global incidence, including a 97% reduction in HAT cases in the former Equateur province of DRC between 2000 and 2012 [5]. However, the screen, diagnose and treat strategy has been unable to effectively control transmission to this level in all endemic foci (e.g. some health zones of Kwilu province, DRC), probably due to insufficient levels of coverage, imperfect diagnostics, or people at high risk of transmission not participating in screening activities.

Where epidemiological and/or control campaign data of infectious diseases are available, data-driven models have proved to be a valuable tool for quantitatively assessing epidemiological assumptions about disease transmission dynamics or evaluating the effectiveness of intervention measures [6–8]. For HAT, data arising from several interventions implemented in recent years have enabled modelling and quantitative analyses of the potential advantages of novel interventions in endemic regions such as Kwilu and former Equateur province in DRC [9–11], Mandoul in Southern Chad [12], and Boffa in Guinea [13]. Nonetheless, many epidemiological aspects of HAT remain unclear, and additional data are needed to fill these knowledge gaps. For example, the role of certain subpopulation groups in maintaining transmission in endemic areas, such as those not covered by screening programmes or at unusually high risk due to behavioral or geographical characteristics; or the potential existence of reservoir animal hosts or asymptomatic human carriers is not fully understood [14].

With the 2030 EOT goal on the horizon, it is crucial to determine which efforts in which locations could maximise the potential benefits of any intervention against HAT. Modelling could provide the HAT community with a better understanding of the important factors affecting observed changes in intensity of disease reporting and explain some of the variations in effectiveness of HAT control and surveillance activities across different settings.

In this study we analyse a longitudinal human epidemiological data set of HAT from former Bandundu province in the DRC to outline how the type of data and its level of aggregation may affect projections of HAT transmission models. Four independent HAT models, fitted to different data aggregation sets, are used to investigate how the level of data aggregation impacts the projections of HAT incidence and likelihood of achieving the EOT goal for current and intensified intervention strategies. Although the 2030 goal is defined as EOT for the continent, and therefore meeting EOT within Bandundu is not directly equivalent, failure to meet the goal in this high-endemicity region would imply failure to meet the global EOT target. Implications of data resolution on the estimated effectiveness of strategy is analysed in order to suggest potential improvements in data collection and availability that could contribute to robust assessment of control programme effectiveness and reliable estimates of HAT elimination.

## Materials and methods

### Data description and assumptions

Former Bandundu province in the DRC has the world’s highest HAT burden despite a significant coordinated effort between national and international HAT control programmes [5]. This province covers an area of 296,500 km^2^ (12.6% of DRC) and accounts for the largest number of cases reported since 2001 in the country (approximately 47.6%).

In this study we used publicly available provincial level human case data from Bandundu province [5] to calibrate models of HAT transmission. The data contains the annual number of positive cases for each stage of the disease detected through active screening and passive detection (the primary HAT control interventions implemented in this area); and the total screened population across the province for the years 2000-2012. Although the geographical scale of this province-level data is large, this data was chosen because - to the authors’ knowledge - this is the only (either publicly or under-request) available data providing details on the stage of reported cases for many consecutive years.

Estimates of the population of Bandundu were taken from publicly available census data [15] for 2000-2012 and a 3% annual growth rate was assumed for projections.

Although target populations are usually estimated prior to each active screening round, this data was not publicly available and the target varies from year to year depending on the health zones screened. To determine a consistent estimate over 13 years, each model assumed a constant proportion of the population at risk over the entire period, either fixed or estimated during model calibration (see details in <u>S2 T</u>ext).

### HAT models

Four independent deterministic models of HAT transmission were used (hereafter named as Model I, Model S, Model W and Model Y) to evaluate the effects of different levels of data aggregation on forward projections.

All of them were based on models previously used in either simulation or data-driven studies [9, 10, 16–18] and include modifications, independently implemented by each group, to improve calibration to the data analysed here. Differences in structural assumptions (e.g. disease progression, heterogeneity in risk to infection) and parameterisation reflect the variety of complexities and biological uncertainties typically found in epidemiological models. Furthermore, a range of different fitting methodologies were employed which also have implications on results. An overview of key aspects of model structure, interventions and fitting procedure is given in Table 1 and more details of each of the models can be found in <u>S2 Text</u>.

**Table 1.**
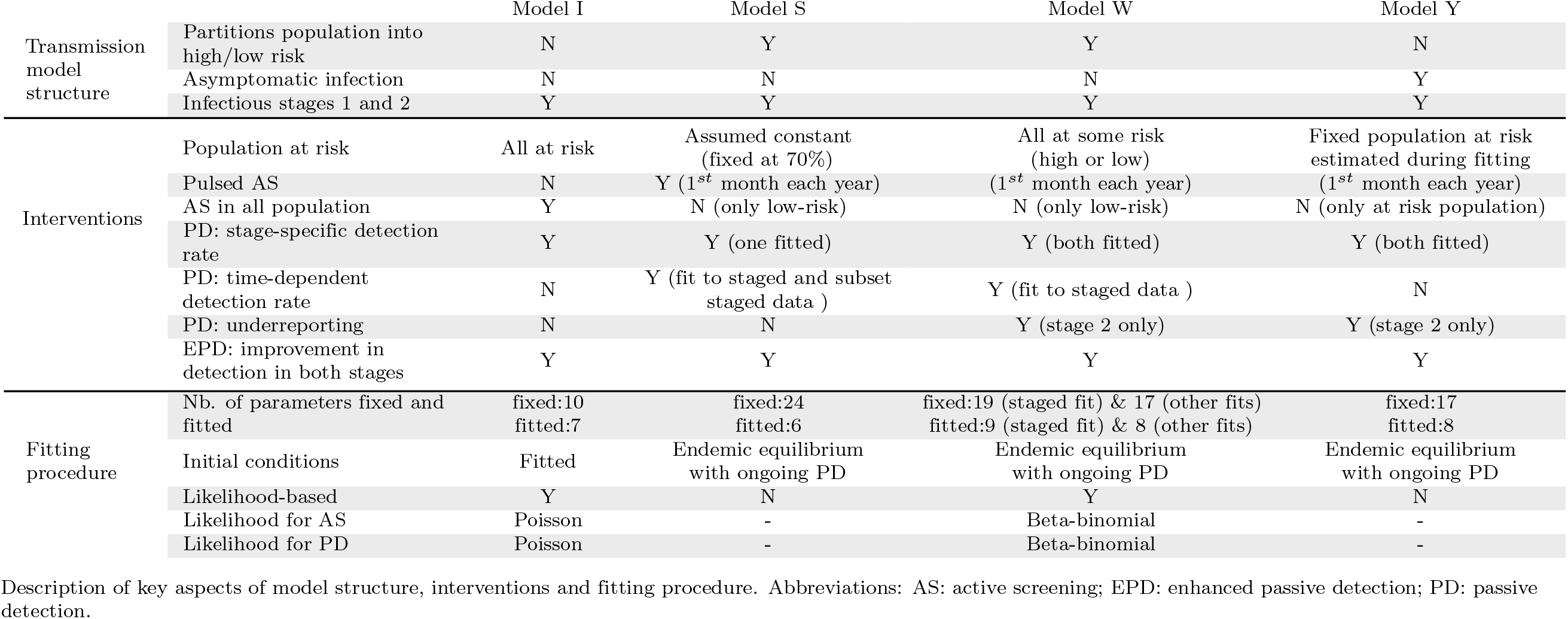
Models overview.

### Model fitting

The reported number of cases detected through active and passive screening and the number of people tested were used to calibrate the models emulating the effects of a typical medical control strategy. The data do not contain information on the timing and duration of active screening, so each modelling group independently managed these aspects (see Table 1).

The models were calibrated to three different configurations of the data to reflect the diversity of data resolution usually available, allowing the analysis of the impact of data detail on both uncertainty and reliability of model projections. The three configurations were labelled: “unstaged data”, “staged data” and “subset staged data”. “Unstaged data” informed the models using the number of HAT cases detected each year (2000-2012), separated by active and passive detection. This type of longitudinal data - where the disease stage is not noted - is typical of data available at smaller administrative levels, such as health zones or health areas in DRC. “Staged data” additionally partitioned the number of cases from the “unstaged data” by disease stage (first or second). The “subset staged data” consisted of a temporal subset of the “staged data”, covering only years 2000-2006. By cutting the data at this point, the improvement observed after 2006 in the detection of stage 1 cases is not yet apparent, and so we expected to see some effects of this in model estimations and projections.

Each group independently chose a calibration method adapted to their own model. The list of fixed parameters used (either obtained from the literature or assumed) and those estimated during the fitting are detailed in the description of each model in <u>S2 Text</u>. Fitting procedures included Bayesian inference using Markov Chain Monte Carlo (MCMC) (Models I and W) and approximate Bayesian computation methods (Models S and Y). In all cases, one thousand samples (i.e. parameter sets) were generated during the fitting step for further estimations and projections. In all cases plots display the median and associated 95% credible interval (CI). For further details on models’ structure, assumptions and fitting procedure, see details in <u>S2 Text</u>.

### Simulated HAT interventions

Four interventions were considered for simulations. They consisted of three medical-based interventions: “active screening”, “passive detection” and “enhanced passive detection”; and “vector control”. A brief description of these interventions is provided below.

- **Active screening (AS)**. This is the screening of the population at large in at-risk locations by mobile teams. Once detected, patients travel to medical centres for treatment. In this study, the reported annual number of people screened was used to estimate the mean active screening coverage. Models that included population heterogeneity in exposure to tsetse (Models S and W) assume that only low-risk people are screened actively.
- **Passive detection (PD)**. This is the diagnosis and treatment of infected people who self-present at medical facilities. HAT models usually assume that passive surveillance detects mainly stage 2 cases, when symptoms are more severe and specific to HAT. The data used in the present work reports a non-negligible proportion of stage 1 cases detected through passive surveillance. For this reason, both stages were assumed to have the potential to be detected in all models.
- **Enhanced passive detection (EPD)**. This is passive screening where the time to detection of infected people is reduced (i.e. improved detection rate per capita). Such improvement could result from one or a combination of changes in current control activities. For example, increasing the number of health facilities (thus increasing the chances of picking cases), mobilising the population at risk or by reducing the time to detection and treatment through improved HAT diagnostic tools including rapid detection tests (RDTs). In DRC, RDTs have been used in many endemic settings between 2013 and 2016 [19, 20], although estimates of the improvement on the associated detection rate have not yet been quantified.
- **Vector control**. This intervention focuses on increasing the mortality and reducing the density of tsetse flies by, for example, deploying insecticidal baits (e.g., insecticidal targets, insecticide-treated cattle) to attract and kill tsetse. In particular, tiny targets [21] offer great promise for the large-scale and cost-effective control of the riverine tsetse species which transmit gambiense HAT [12, 21–23]. Tiny targets were first deployed in DRC in 2015, in Yasa Bonga health zone, and they are currently being used in three health zones of former Bandundu province.

With these four interventions, three different strategies were investigated that reflect either the current control and surveillance programmes or strengthened strategies to accelerate the elimination of HAT. These are:

- Strategy 1: also referred to as “baseline”, this strategy represents the standard control method in Bandundu consisting of continuing active screening and passive detection at present rates.
- Strategy 2: consists of vector control in addition to the baseline strategy, as is currently being implemented in Yasa-Bonga, Masi Manimba and Kwamouth health zones of Bandundu. In the models, vector control was assumed to reduce tsetse populations by 60% after one year, which is a conservative estimate from intervention trials conducted in other HAT foci [12, 21, 22].
- Strategy 3: assumes enhanced passive detection, in addition to the annual active screening campaign. For this strategy, Models I and S doubled the passive detection rate of both stages while models explicitly including underreporting (Models W and Y) assumed both a doubled passive detection rate and halving of underreporting. We also assumed that the treatment rate of detected cases remained the same so that increased detection led to a corresponding increase in the treatment rate.

The calibrated models were used to simulate the “future” effects of these three strategies (Table 2) in order to compare, for each model, the effects of the different types of data aggregation used for calibration, on projections and associated uncertainty under different control strategies. In all cases the baseline strategy matched the period corresponding to the data, and assumed a continuation of standard passive surveillance and past mean active screening levels informed by the data for projections into the future.

**Table 2.** Different types of future strategies considered in model projections.

| Strategy | Interventions |  |  |
| --- | --- | --- | --- |
|  | Passive | Active | Vector control |
| 1 | Standard | Mean of historic data | - |
| 2 | Standard | Mean of historic data | 60% reduction |
| 3 | Enhanced | Mean of historic data | - |

Model simulations estimated *(i)* annual stage-specific cases reported from both active and passive screening; *(ii)* new transmissions by year; and *(iii)* year of EOT

## Results

### Model fits

#### Reported cases

Fig 1 shows the data from 2000 to 2012 of the total reported HAT cases in Bandundu and the calibrated simulations of the four models to three different data configurations (median with the 95% credible interval (CI)) under the “baseline” control strategy. All fits of all models consistently reproduced the decreasing trend observed in data.

**Fig 1.**
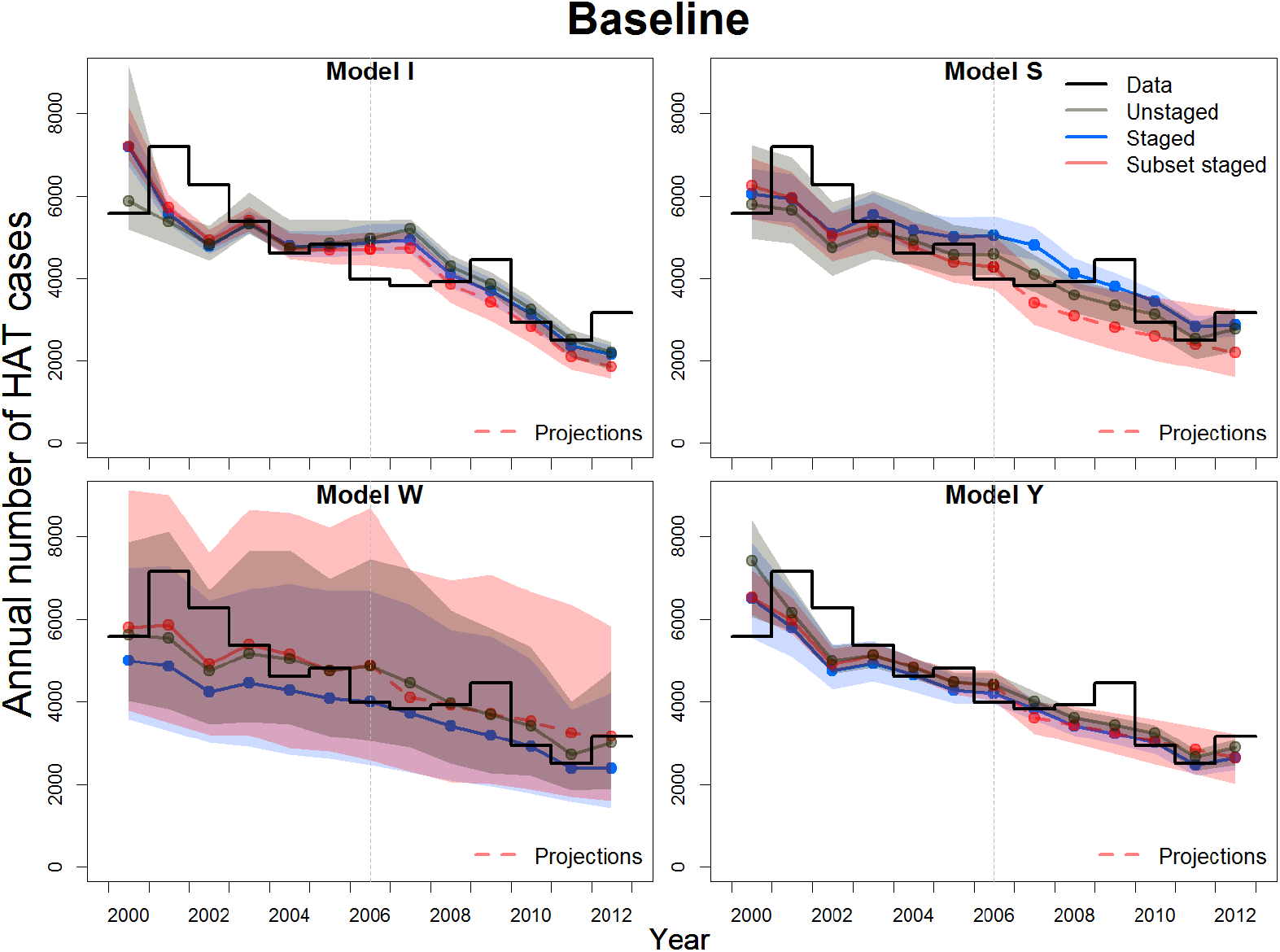
Former Bandundu province reported data and estimated reported cases. Estimated reported cases from model calibrations to three different configurations of the data for a baseline strategy composed of annual, pulsed active screening and continuous passive detection. The median (as a point) and the corresponding 95% CI (shaded region of the same color) are shown in each case. Dashed lines indicate projections from the fit to the subset staged data.

However for most model fits, the 95% CI did not cover all the data points in time series, as is often the case for peaky stage-specific data dominated by a decreasing trend (Fig S1.1 in <u>S1 T</u>ext). Models provided varying levels of uncertainty, mainly explained by differences in fitting methods as well as model structure and parameterisations. Despite all these differences, the fit to the longer, staged data set generated less uncertainty in all four models, with worse and varying performance for the fits to the other data sets.

While for Model W the medians from the fit to staged data gave the lowest estimation compared to the other two fits, for Model S this trend was the opposite for most years. For Models I and Y such a clear trend was not observed among medians.

#### Proportion of stage 1 cases

The increasing trend in the proportion of stage 1 cases out of total reported cases across years (Fig 2) indicates improved screening in Bandundu; this is observed in both active and passive case data <u>(S1 Fig</u> and <u>S2</u> Fig). Model fits not informed with staging ratios produced the worst estimates of this proportion and the highest uncertainties (Fig 2), reflecting a wide range of possible configurations of the proportion of stage 1 infections compatible with such unstaged data either in active screening <u>(S1</u> Fig), passive detection <u>(S2</u> Fig), or both.

**Fig 2.**
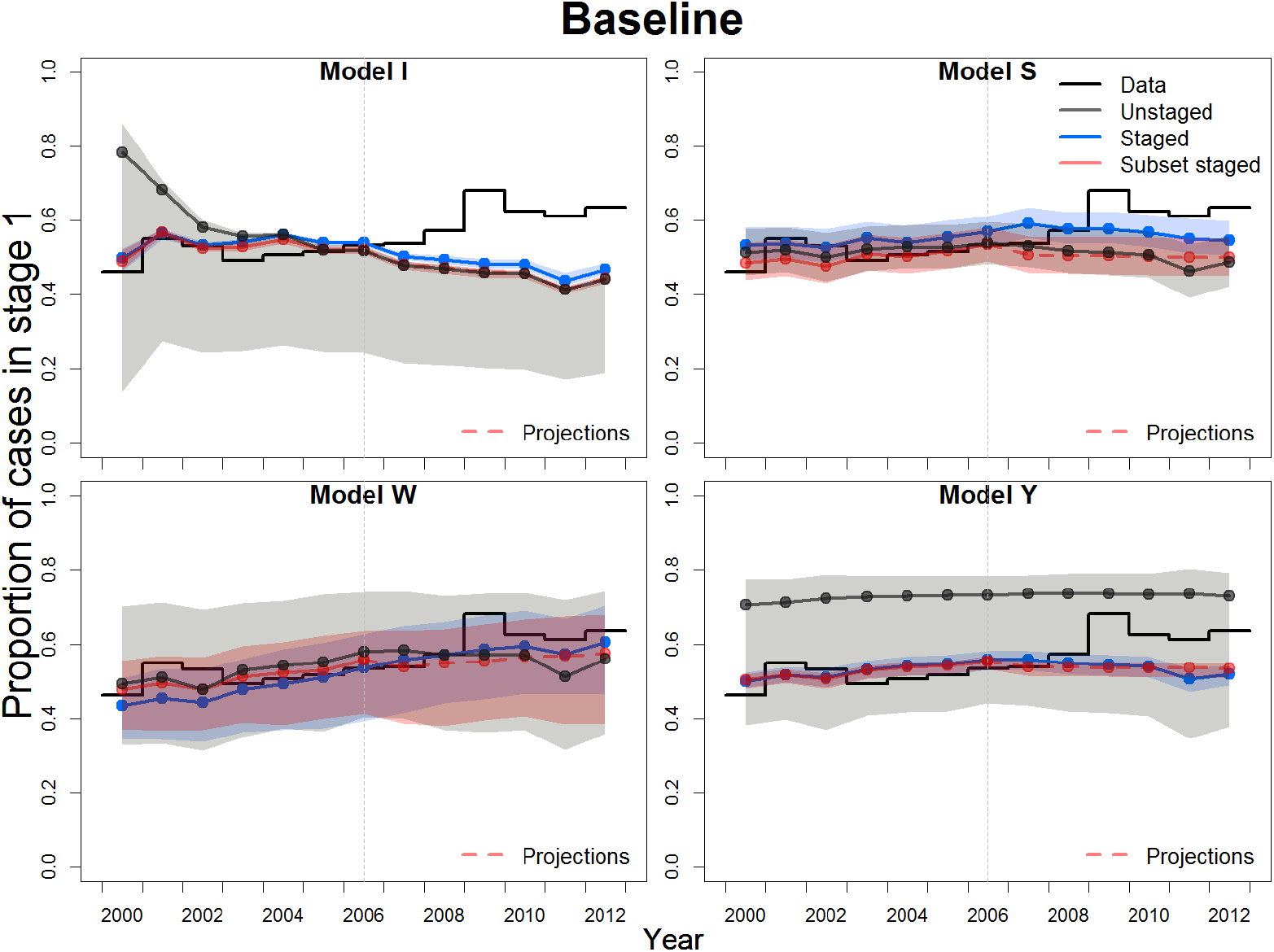
Proportion of stage 1 cases. The estimates for the four models fitted to three different configurations of the data under the baseline strategy are shown. The posterior median is shown as a point and 95% CIs shaded. Dashed lines indicate projections from the fit to the subset staged data.

The variety of assumptions in the models about intervention implementation, including how annual active screening was applied (continuous vs. pulsed, one vs. several per year) or which proportion of Bandundu province population was assumed to be at risk of infection (Table 1), explain in part the variety of results in the proportion of stage 1 cases for different fits. Model W fitted to the full staged data was the only model that reproduced the increasing trend in active screening <u>(S1</u> Fig); and only Models S and W, which assumed an improvement in passive detection rate, reproduced the increasing trend in passive detection, with systematic overestimation in Model S (S<u>2 Fig)</u>. For these two models, it is clear how the fit to the subset staged data, where the improvement in the passive detection is not yet apparent (contrary to the fit to the full staged data), conditions the models to project lower ratios of stage 1 to stage 2 cases from 2007 onwards.

### Projections for future case reporting and transmission

Model projections under all fits came to a consensus that continuing the baseline medical strategy would lead to a sustained but slow reduction of the annual incidence; however some simulations of Model S (86 out of 1000) fitted to unstaged data suggested transmission would increase under baseline strategy (Figs A-D in <u>S3 T</u>ext). The latter is an example of how some parameters sets, although overall can reproduce unstaged data trends, can have an underlying epidemiology promoting increasing transmission despite continued active screening and passive detection levels. Note that these scenarios are not observed when Model S is fitted to the more informative staged data that impose further constraints to the posterior parameter distributions.

As expected, the models indicated that improved or complementary interventions would accelerate this path towards reduced incidence <u>(S3 T</u>ext). Notably the longer staged data set produced the least uncertainty in all models for projections on annual incidence (Figs A-D in <u>S3 Text)</u> and associated reported cases (Figs A-D in <u>S4 T</u>ext). Assuming that projections under the staged data are most robust, the unstaged data generated systematic overestimation in transmission and associated report case projections for any strategy considered in three models (Models S, W and Y); for Model I, a slight discrepancy in projections of new cases was observed, although values from both fits were close and overlapped in projections of reported cases. Model I generated the most optimistic scenarios, with a relatively homogeneous range of projections for the different fits and small uncertainties compared to the other models, with and values on the order of *≈*100 new detected cases or fewer by 2030 for Bandundu province.

Table 3 presents the proportion of simulations (i.e. realisations of different parameter sets) for different fits and models where the 2030 zero transmission goal was achieved, and provides an alternative view on how adding or removing relevant data impacts the models’ projections under different control strategies explored. Here “elimination” is defined as *<*1 transmission case per million individuals per year as in previous work using these deterministic models [18].

**Table 3.** Probability of different strategies achieving elimination by 2030. EOT is defined in the models as *<*1 new transmission per 1,000,000 people. In each case simulations of 1000 parameter sets were used.

| Fit | Strategy |  |  | Model |
| --- | --- | --- | --- | --- |
|  | Baseline | Vector control | EPD |  |
| Unstaged | 0.167 | 1 | 0.167 | I |
| Staged | 0 | 0.656 | 0 |  |
| Subset staged | 0 | 1 | 0 |  |
| Unstaged | 0 | 0.206 | 0 | S |
| Staged | 0 | 0.551 | 0 |  |
| Subset staged | 0 | 0.836 | 0 |  |
| Unstaged | 0 | 1 | 0 | W |
| Staged | 0 | 1 | 0.984 |  |
| Subset staged | 0 | 1 | 0 |  |
| Unstaged | 0 | 1 | 0 | Y |
| Staged | 0 | 1 | 0 |  |
| Subset staged | 0 | 1 | 0 |  |

In all but one case (Model I fitted to unstaged data), the models found that it was extremely unlikely that elimination would occur by 2030 using the baseline strategy. All fits for Models W and Y predicted elimination using vector control tools in addition to the baseline strategy. The least optimistic predictions were observed in Model S, in accordance with higher values and slower reduction in transmission projections when compared to other models’ predictions (Fig B in <u>S3 T</u>ext). For Model I, the fit using the staged data set showed less optimistic predictions, which is consistent with the transmission projections generated by each fit under this model (Fig B in <u>S3 T</u>ext). Only two models under different fits (Model I fitted to unstaged data and Model W fitted to staged data) showed that elimination was possible for enhanced passive detection (167 and 984 out of 1000 samples, respectively).

For a weaker definition (*<*1 transmission case per 100,000 individuals per year), only Model I suggested elimination could be achieved for the baseline strategy, and all model-fit combinations agreed on vector control achieving elimination by 2030.

Substantial improvement in elimination probabilities under enhanced passive detection in Models I and W contrasted to results of Models S and Y where no significant changes were found <u>(S1 Table)</u>. The higher disparity among models in predicting elimination probabilities under enhanced passive detection reflects the influence of structural assumptions, in both HAT transmission dynamics but also in modeling control activities that can lead to such different projections.

## Discussion

A suite of independent mathematical models of HAT transmission were calibrated to publicly available data from Bandundu province, DRC, to evaluate the effects of different levels of data aggregation on model performance and projections under current and improved control strategies.

### Informing staging data

Distinguishing cases by stage is inherent to HAT epidemiology due to the way treatment is currently administered. The results here showcase the impact that neglecting staging information in data reporting has on subsequent model estimates and predictions.

Although similar patterns of annual incidence can be obtained from models calibrated to unstaged and staged data, the underlying HAT dynamics for such similar incidence patterns can differ strongly (as indicated by the proportion of stage 1 cases detected), affecting any inference or projection on transmission risk. Contrasting projections between staged and unstaged fits demonstrate how this aspect of HAT epidemiology can impact our optimism about a particular strategy. A key example is that model calibrations using staged data for Bandundu province strongly suggest that passive detection rates have improved over time, whilst this is unobservable in the unstaged data.

The data that countries use to determine their elimination policies for HAT are usually limited and come mainly from screening activities. Our results emphasize the need for incorporating staging information in data sets. With current screening protocols, minimal additional effort in data recording is required to systematically include staging, which would help to reduce uncertainties in assessing progress towards elimination goals.

In the future, staging information may no longer be collected if new diagnostic tools and treatments are stage-independent. For example, the new drug, fexinidazole [24], is an all-in-one oral treatment for both stages recently approved by the European Medicines Agency. However, until such tools become part of regularly implemented policy, we emphasise the utility of making routinely collected staging data available. Furthermore, if records of historically collected staging data exist, making these available would substantially improve the reliability and predictive capability of mathematical models.

### Time scales and informing on time surveys of active screening

Over half of the total number of stage 1 cases reported between 2000 and 2012 come from active screening. In general, as in this study, data is annually aggregated and so the timing and the duration of active campaigns is unknown. As with current staging data, this information is recorded at lower administrative levels, but is often lost in higher level data sets. Systematically adding temporal data to current routine data collection and collation would enable exploring a variety of case-specific time related epidemiological factors such as the optimal frequency of interventions for achieving specific local goals.

### Data delays

There are routinely delays between case detection in the field and the availability of the data for modeling purposes. The extreme example of a six years delay between data collection and availability considered in this study, though unlikely due to improvements in data availability, is chosen to demonstrate how the absence of up-to-date data impacts model predictions. One or two missing years would still provide less accurate results than up-to-date data, especially due to the lack of information on recent active screenings. Nevertheless, we expect that model predictions generated with fewer missing years would generate predictions more similar to predictions using the full data set than those generated with six missing years as investigated in this study.

As we approach elimination, including recent data sets is necessary to better assess the actual trends, as our results have suggested. Use of most recent data sets can be sufficient to reproduce current epidemiological trends and the absence of these data sets could affect model projections, especially for short timelines. Improvements in the time between data collection and availability could enable modelling to provide more up-to-date guidance and monitor for early-warning signs of obstacles on the road to elimination.

### Province level data vs health zone level data

Aggregated province-level data for endemic HAT regions lose information on the geospatial variation of HAT incidence and screening coverage at lower administration levels that are more compatible with the epidemiological scale of HAT transmission and control. This may explain why although all model fits could capture the decreasing trend in the number of reported cases, they could not reproduce certain peaks observed in stage 1 cases (in 2002 and 2009) from active screening. The models assumed a fixed, spatially homogeneous risk of transmission in Bandundu province, even though large differences between central and southern health zones of Bandundu province had been estimated for this period [5]. Model W uses overdispersion parameters to capture the variation in data between different years, so fitting to finer resolution data would likely explain the source of this variation, and reduce the very large credible intervals from the current results.

The peaks observed in the data could arise due to differences in HAT prevalence in the geographical areas in which the active screening occurs between years, due to differences in the quality or coverage of the screening campaigns between years, or reflect true inter-annual variation in HAT epidemiology. Only detailed case data at a finer spatial scale could help models to explore alternative assumptions, capture spatial heterogeneity to better identify geographic reservoirs and improve predictions in global HAT status. Model calibrations at a health zone or finer spatial scale are needed to directly guide practical strategy planning at a local level. The WHO HAT Atlas is one such valuable source of geolocated data in DRC (available upon request from the WHO); and although staging information is typically not available for cases before 2015, recent entries are staged.

### Complementary interventions to meet elimination goals

Projections suggest that, at the province level, the continuation of traditional active and passive screening is unlikely to be sufficient to attain EOT by 2030 across most models and fits. The groups therefore simulated other complementary strategies which built upon these baseline interventions to examine if any were sufficient to achieve this goal.

#### Vector control

Our results agree with previous modelling work indicating that potential strategies that integrate vector control with medical interventions could accelerate progress towards elimination, particularly in high endemicity or persistent hotspots [10, 11, 13, 17, 18]. This is consistent with reductions in HAT transmission reported after implementation of cost-effective vector control methods in highly endemic locations in Guinea [22] and Chad [12].

Although integrating vector control with current medical interventions at large spatial scales such as Bandundu province (around 296,000 km^2^) may not be operationally feasible, extending tsetse control interventions to active foci of HAT transmission is feasible and likely to be efficient, particularly as transmission decreases and programmes reduce screening activities. Vector control is currently being implemented in hotspots in Bandundu (totalling approximately 3000 km^2^) and in the West Nile region of Uganda (covering approximately 5000 km^2^). Regularly updated epidemiological and entomological data from areas that have added this intervention to HAT screening activities would facilitate the analysis of progress towards elimination objectives, and provide an indication of protection against infection due to vector control.

Additionally, secular changes, such as socio-economic development, urbanisation and changes in land use, would likely lead to sustainable reductions in tsetse population densities and consequently in HAT transmission, similarly to what has been reported for other vector-borne diseases [25]. The impact of such secular changes was not addressed in this study but will become more important as transmission reduces further.

#### Enhanced passive detection

This study found that, for passive detection, the increase in the ratio of stage 1 to stage 2 cases from 2006 onwards is an indicator of an already improving passive screening system in this part of DRC. Although this is to be expected considering the increased disease control efforts in the region, it is the first time that the improvement in the passive detection rate has been quantified in a mechanistic modelling framework.

Furthermore, this trend is not observed in other former provinces of DRC for data from the same period [5].

An improvement in time to detection is likely to have been driven by a combination of causes, including improvements in access to care from increased awareness by the population at risk and an increase in the number of health facilities; and improvements in diagnostic tools including the use of digital technologies and RDTs (FIND 2016, [20, 26]). Moreover, new “test-and-treat” strategies combining RDTs with fexinidazole could lead to earlier and more cases treated.

Although our results suggest that enhanced passive detection could not be sufficient to achieve short-term reduction goals, its associated sustained effect on reducing transmission, projected by all models, indicates this strategy should be considered for areas in Bandundu where past activities did not reduce HAT transmission as expected.

#### Reactive screening

As the number of reported cases decreases, reactive case detection, i.e., deploying active screening in a given area following detection of a case by passive screening, may be a potential cost-effective strategy. Such a complementary strategy has already been implemented in some regions of Uganda, Chad, Kongo Central and Angola. The inherent spatial aspect of reactive screening implies that modelling elimination would benefit greatly from geolocated and timed case data from different settings. This would allow for an improved assessment of spatially-related measures of HAT transmission risk to inform the appropriate targeting of interventions in space and time to achieve elimination and prevent resurgence.

### Cost implications

Naturally each of the different strategies will affect the total cost of HAT interventions in the region, with complementary strategies costing more than the baseline in the short-term due to the extra resources used. Strategies which cost more in the short-term could result in earlier EOT, and therefore may lead to earlier cessation of active screening interventions compared to baseline. This could yield lower long-term costs, but it is non-trivial to assess the costs of these interventions without simulating cessation strategies and using a cost model.

Cost-effectiveness analyses using dynamic modelling frameworks require assessment of health outcomes (such as years of life lost, and disability adjusted life years due to disease) against a budget or willingness-to-pay threshold which can lead to strategies which are not the least expensive being selected due to the relative gain in health benefits [27]. This health-economic work is beyond the scope of the present study, which primarily seeks to address the impact of data aggregation on model fitting and projections. Assessment of cost-effectiveness is clearly an interesting and important objective for future analyses which aim to provide specific, regional recommendations for strategy selection. Such work would ideally provide more local strategy guidance (smaller than the province scale considered here) so that only regions that require complementary interventions include them rather than assuming blanket coverage of additional strategies across large areas.

### Extrapolations to other aspects of data

Between 2011 and 2013, a study was performed to analyse the effects of coordinated vector control (using tiny targets) and mass screening in an area of over 300 km^2^ in the endemic focus of Boffa in Guinea [22]. This study recorded highly detailed pre-intervention geo-referenced data of households and inhabitants (familial clustering via a unique code; name, sex and age of family members); annual screening data; and vector and vector control data (15 targets/km^2^, estimates of initial tsetse fly densities, trap location, survey duration); as well as subsequent updates including new families and seasonal workers. Although such a comprehensive and rich data set can provide a much deeper understanding of HAT epidemiology and the quantitative impacts of control interventions on transmission, scaling up such studies to cover larger areas is likely to be too costly to be feasible. A potential alternative would be to enrich current standard data collection/collation from screening activities with questionnaires providing additional demographic information on infected individuals (e.g. age, gender, occupation, characteristics of house location) to better assess people at risk, their participation in screening and their impact on transmission. Although this too may be costly in higher transmission areas, it may be feasible close to elimination, where case numbers are low and such enriched data would be particularly useful in identifying potential new cases, as programmes move from untargeted active surveillance to reactive strategies.

Table 4 summarises different, but not exhaustive, data which, if available, could be used in modelling studies to identify potential beneficial adjustments in future activities and to develop new frameworks for evaluating the path towards elimination and post-elimination scenarios.

**Table 4.**
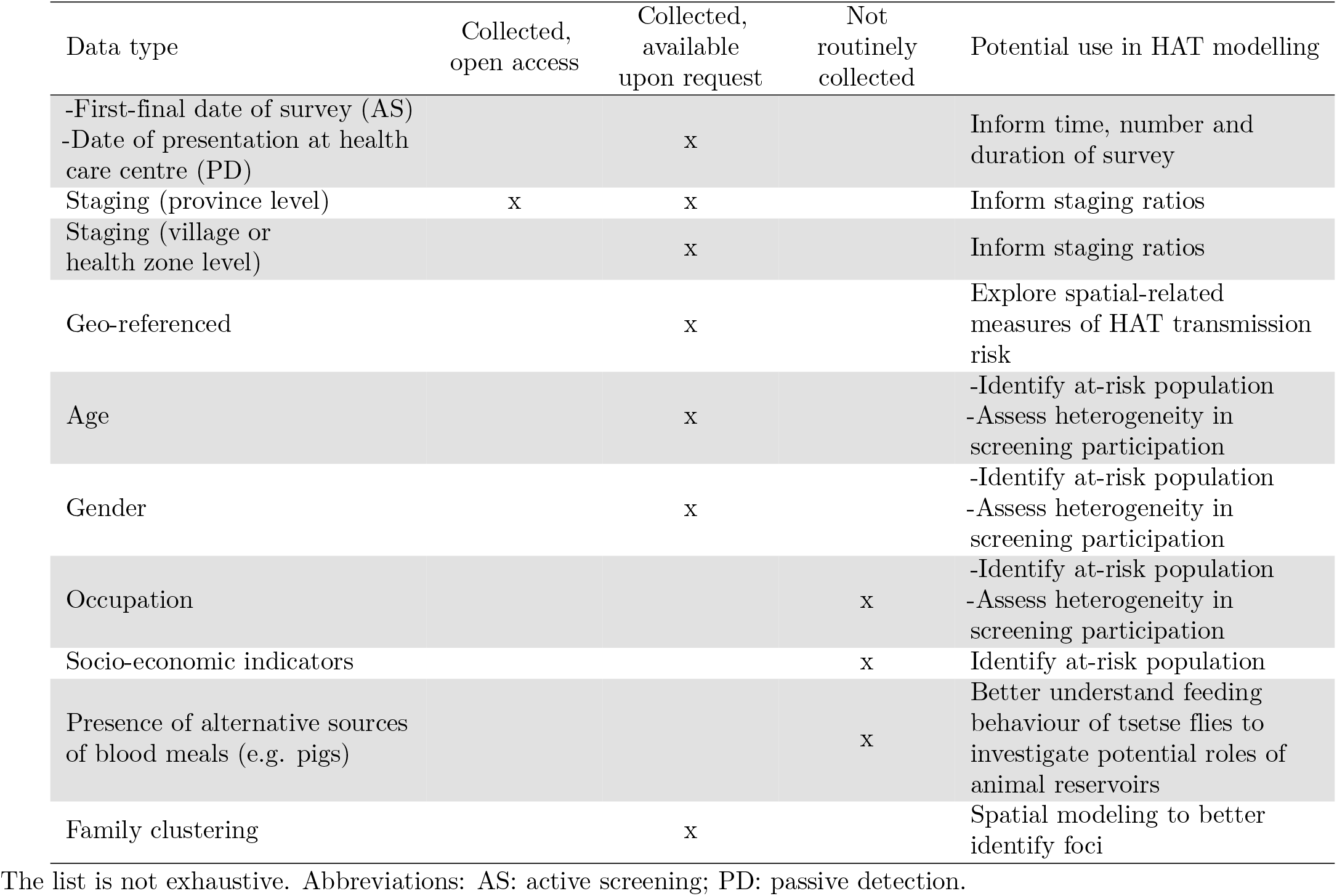
Summary of relevant data and its potential use in HAT modelling.

### Conclusions

We investigated the role of the type and level of aggregation of epidemiological data on recommended control strategy by analysing publicly available HAT case data using four different mathematical models. Our results show that the lack of detailed epidemiological information, such as missing staging or truncated time series data, impacts model recommendations for strategy choice: it can increase our prediction intervals and either over or underestimate effectiveness of interventions. Across all models and configurations of data sets, the present study suggests that adding vector control to current active and passive screening is likely to be the best strategy to reduce transmission quickly in this region (former Bandundu province, DRC). For the other strategies (including current active and passive detection, and enhanced passive detection with active screening), the probability of achieving elimination and the prediction of the time to elimination vary among models and depend on the data configuration used for calibration.

Our study suggests that improved availability of epidemiological data, particularly longer time series which include recent data and information on disease stage, would reduce uncertainties in the prediction of future HAT dynamics. In particular, staging data allow a better estimate of the improvements made in passive detection, and subsequent reduction in HAT transmission. Given the highly focal nature of HAT, we expect that models fitted to recent staged data at smaller spatial scales (e.g. health zone level) will provide valuable information for local planning, monitoring and adapting HAT interventions to reduce transmission and achieve elimination.

## Data Availability

All data is available in the supplementary information.

## Supporting information

**S1 Text. Remarks on former Bandundu province case report data. S2 Text. Model descriptions**.

**S3 Text. Projections on new infections**. Projections on the annual incidence of new infections for all combinations of models and data sets.

**S4 Text. Projections on case reporting**. Projections on the annual HAT cases for all combinations of models and data sets.

**S1 Table. Probability of elimination (zero transmission) by 2030 with a weaker threshold**.

**S1 Fig. Stage 1 reporting in active screening**. Proportion of stage 1 to total cases reported from active screening, and the corresponding estimation for a baseline strategy under different fitting. The posterior median is shown as a point. Dashed lines indicate projections based on fit to subset staged data.

**S2 Fig. Stage 1 reporting in passive detection**. Proportion of stage 1 to total cases reported from passive detection, and the corresponding estimation for a baseline strategy under different fitting. The posterior median is shown as a point. Dashed lines indicate projections based on fit to subset staged data.

## Acknowledgements

The authors thank WHO HAT team for facilitating access to the HAT Atlas data; and to José R. Franco and Gerardo Priotto for helpful discussion and comments on this manuscript. Calculations for the Model S were performed at the sciCORE (http://scicore.unibas.ch/) scientific computing core facility at the University of Basel.

